# DNA methylation patterns and disease activity in a longitudinal cohort of lupus patients

**DOI:** 10.1101/2020.09.18.20197525

**Authors:** Patrick Coit, Lourdes Ortiz-Fernandez, Emily E. Lewis, W. Joseph McCune, Kathleen Maksimowicz-McKinnon, Amr H. Sawalha

**Author notes:** Please address correspondence to Amr H. Sawalha, MD. Address: 7123 Rangos Research Center, 4401 Penn Avenue, Pittsburgh, PA 15224, USA. **Conflict of interest:** The authors have declared that no conflict of interest exists.

## Abstract

Epigenetic dysregulation is implicated in the pathogenesis of lupus. We performed a longitudinal analysis to assess changes in DNA methylation in lupus granulocytes over 4 years of follow up and across disease activity levels using 229 patient samples. We demonstrate that DNA methylation profiles in lupus are influenced by ancestry-specific genetic variants and are highly stable over time. DNA methylation levels in two CpG sites correlated significantly with changes in lupus disease activity. Progressive demethylation in *SNX18* was observed with increasing disease activity in African-American patients. Importantly, demethylation of a CpG site located within *GALNT18* was associated with the development of active lupus nephritis. Differentially methylated genes between African-American and European-American lupus patients include type I interferon-response genes such as *IRF7* and *IFI44*, and genes related to the NFkB pathway. *TREML4*, which plays a vital role in toll-like receptor signaling, was hypomethylated in African-American patients and demonstrated a strong *cis-*meQTL association.

## Introduction

Systemic lupus erythematosus (SLE or lupus) is an autoimmune disease of incompletely understood etiology. Genetic, epigenetic, and environmental factors are thought to play key roles in the immune dysregulation underlying the development of the disease(1). Lupus is characterized by the production of autoantibodies to nuclear antigens, and a remitting-relapsing disease course that can target multiple organ systems(2). Frequent disease flares and prolonged periods of active disease are associated with a more deleterious outcome in lupus patients and a higher risk of tissue and organ damage(3).

Lupus is associated with changes in gene expression, including prominent type I interferon and neutrophil gene signatures in the peripheral blood(4-7). Further, increased disease activity in lupus is associated with transcriptional profiles implicating different innate and adaptive peripheral immune cells in individual patients followed longitudinally(7). Notably, progression to active nephritis in lupus patients was associated with gradual enrichment in neutrophil transcripts(7). Indeed, a prominent role for neutrophils in the pathogenesis of lupus is being more clearly elucidated(8).

DNA methylation, an epigenetic mechanism that regulates gene expression, is altered in the immune cells of lupus patients and is potentially influenced by both environmental and genetic factors(9). DNA methylation defects in lupus are suggested to promote an overactive immune response when exposed to inflammatory signals like autoantibody-autoantigen complexes or endogenous nucleic acids(10-12). Methylation quantitative trait loci (meQTLs) are genetic polymorphisms that are associated with DNA methylation either directly through alteration of CpG dinucleotides or at a distance through an intermediary process. MeQTLs identified in prior lupus studies show enrichment for lupus susceptibility genes and type I interferon response genes suggesting that altering DNA methylation levels at specific loci could be a potential mechanism by which risk alleles contribute to disease susceptibility in lupus(13-15). Lupus susceptibility is significantly higher in patients of non-European ancestry, who are also more likely to develop more severe disease even after accounting for the influence of social and environmental factors(16). Thus, meQTL analysis provides a potential approach to better understand the mechanisms underlying the observed differences in disease manifestations and outcomes in lupus patients of different ancestries.

Recent work investigating DNA methylation changes in lupus and associated downstream effects and underlying upstream regulatory mechanisms have resulted in significant insights into the pathogenesis of lupus and the identification of novel therapeutic targets for the disease. Further, DNA methylation changes have been suggested as diagnostic markers and markers that can potentially predict specific disease manifestations in lupus(11, 17, 18). However, DNA methylation studies in lupus to date have been cross-sectional, and longitudinal studies investigating epigenetic changes in patients with lupus over time have not been reported.

We have previously demonstrated robust demethylation of interferon-regulated genes in lupus neutrophils compared to normal healthy controls(12). In this study, we investigate neutrophil DNA methylation changes over time and across disease activity levels in a cohort of lupus patients followed longitudinally for up to about 4 years. Moreover, we sought to increase our understanding of how DNA methylation is impacted by the genetic background. We compared DNA methylation patterns between African-American and European-American lupus patients, performed meQTL analyses in lupus neutrophils, and identified CpG sites that show methylation changes correlating with disease activity and the development of lupus nephritis across the course of the disease.

## Methods

### Study Participants and Demographics

54 female lupus patients were recruited from the University of Michigan Health System and Henry Ford Health System for this study (**Supplementary Table 1**). Our cohort included 32 patients of European-American ancestry and 22 patients of African-American ancestry. Patients were followed over a 43 month period. The patients selected for this study had at least one change in disease activity as measured by the systemic lupus erythematosus disease activity index (SLEDAI) score across all timepoints. This resulted in a total of 229 timepoints across all patients (4 median timepoints per patient; range: 2-11 timepoints). The mean age of patients at the initial visit was 41.0±13.1 years (mean±sd; range: 19-70 years). The mean SLEDAI score of patients was 3.9±3.9 (mean±sd; range: 0-20) at their initial visit and 4.0±3.7 (mean±sd; range: 0-20) across all timepoints (**Supplementary Figure 1**).

**Table 1:** Correlation of DNA methylation and disease activity in a cohort of African-American lupus patients (n = 22) across 93 sample after adjusting for age using a mixed effects model. All sites represented here met our suggestive significance threshold of P < 0.10 and bolded P-values meet our significance threshold of P < 0.05. F-value (Satterthwaite) is the F-value for the F-test conducted by lmerTest using the Satterthwaite method for denominator degrees of freedom.

| African-American Patients (n = 22; 93 timepoints) |  |  |  |  |  |  |
| --- | --- | --- | --- | --- | --- | --- |
| Probe ID | Genes | Location (hg38) | SLEDAI<br>Score<br>Coefficient | F<br>(Satterthwaite) | P-value | FDR-adjusted P-Values |
| cg26104306 | *SNX18* | chr5:54517014 | -0.046 | 36.57 | 4.61E-08 | **3.38E-02** |
| cg06708913 | - | chr12:89880778 | 0.055 | 34.77 | 9.35E-08 | **3.43E-02** |
| cg24682077 | *FGD1* | chrX:54496205 | -0.026 | 30.33 | 3.72E-07 | 5.35E-02 |
| cg15563677 | *ELMSAN1;RP5-1021I20.1* | chr14:73788514 | -0.039 | 31.15 | 3.73E-07 | 5.35E-02 |
| cg26138978 | *EFNB2* | chr13:106530743 | -0.038 | 30.14 | 3.77E-07 | 5.35E-02 |
| cg17038326 | - | chr3:27614709 | -0.049 | 30.64 | 4.38E-07 | 5.35E-02 |
| cg22284518 | - | chr16:49351267 | -0.016 | 29.63 | 5.90E-07 | 6.18E-02 |
| cg00465267 | *NFATC2* | chr20:51497407 | 0.038 | 29.28 | 6.76E-07 | 6.19E-02 |

All patients in this study fulfilled the American College of Rheumatology classification criteria for systemic lupus erythematosus(19). The institutional review boards of each institution approved this study and all patients signed informed consent prior to study enrollment.

### DNA Isolation

Whole blood was collected from each patient at each time point during clinic visits in vials containing EDTA. Granulocyte fractions were isolated using density centrifugation with Ficoll-Histopaque (GE Healthcare, Chicago, IL, USA). Genomic DNA was isolated from the enriched granulocyte layer using either phenol-chloroform extraction or Qiagen DNEasy Blood and Tissue kit (Qiagen, Germantown, MD, USA), or following the removal of red blood cells using dextran (Sigma-Aldrich, St. Lois, MS, USA) and hypotonic lysis(20). DNA was eluted in water and quantified using Qubit DNA fluorescence quantification assays (Thermo-Fisher, Waltham, MA, USA).

### DNA Methylation Measurement

350ng of DNA from each sample was bisulfite converted using the EZ-96 DNA Methylation Kit (Zymo, Irvine, CA, USA) following the manufacturer’s instructions. Samples were hybridized to the Infinium MethylationEPIC array (Illumina, San Diego, CA, USA) to asses site-specific DNA methylation of over 850,000 methylation sites across the genome. Samples were randomized across all arrays to minimize batch effects. Sample hybridization and array scanning were performed at the University of Michigan Advanced Genomics Core.

### DNA Methylation Quality Control and Analysis

DNA methylation data analysis was performed in the R statistical computing environment (v.3.6.3)(21). Raw .idat files were generated for each sample and read into the R package *minfi* (v.1.32.0) for quality control and downstream analysis(22, 23). Probes with less than three beads and zero intensity values across all samples were removed according to best practices as implemented by the *DNAmArray* package (v.0.1.1)(24). Then, background signal and dye bias were corrected followed by normalization of signal intensities using functional normalization in the *preprocessFunnorm*.*DNAmArray* function(24, 25). This method uses the first three principal component values calculated from signal intensities of control probes present on all array spots to correct for technical variation. Probes with detection P-values < 0.01 were removed as were probes that returned signal intensities in fewer than 98% of samples. Signal intensities were then converted to M-values with a maximum bound of ±16. M-values were used for all regression testing and converted to beta values (0-100% methylation scale) using *minfi* for reporting.

We masked any probes with potential technical issues if the probe met any one of the following criteria described by *Zhou, Laird & Shen (2017)*(26): A unique probe sequence of less than 30bp, mapping to multiple sites in the genome, polymorphisms that cause a color channel switching in type I probes, inconsistencies in specified reporter color channel and extension base, mapping to the Y chromosome, and/or having a polymorphism within 5bp of the 3’ end of the probe with a minor allele frequency (MAF) > 1% with exception of CpG-SNPs with C>T polymorphisms which we retained for analysis. Batch correction was performed using the *ComBat* function in the *sva* (v.3.34.0) package(27).

We implemented a mixed correspondence analysis with the *PCAmixdata* package (v.3.1) to calculate eigenvalues using patient medication data for prednisone, hydroxychloroquine, azathioprine, mycophenolate mofetil, and cyclophosphamide(28). The top four components accounted for a cumulative 88.4% of variability in the medication data. Each component value was used as an independent variable in regression analysis to adjust for medication usage across individuals.

### Genotyping and Methylation Quantitative Trait Loci (meQTL) Analysis

Genotyping data were generated using Infinium Global Screening Array-24 v2.0 (Illumina, San Diego, CA, USA) according to the manufacturer’s instructions. Stringent quality controls (QC) were applied before analyses using *PLINK* (v.1.9)(29). Single nucleotide polymorphisms (SNPs) with a genotyping call rate < 98%, MAF < 5%, and deviating from Hardy-Weinberg equilibrium (HWE; P-value < 1E-03) were filtered out. Samples were removed if they had a genotyping call rate < 95%. Sex chromosomes were not analyzed. About 100,000 independent SNPs were pruned and used to perform principal component analysis (PCA) with *Eigensoft* (v.6.1.4) software (**Supplementary Figure 2**)(30). Genotyping data of a single African-American lupus patient was removed at the QC step due to failing quality measures. All meQTL analyses presented in this paper are obtained from the methylation and genotyping profiles of n = 21 African-American and n = 32 European-American lupus patients.

### Gene Set Enrichment Analysis

Gene annotation of CpG probes was done using GENCODE v22 (hg38) annotations from a manifest file produced by *Zhou, Laird & Shen (2017)*(26). Gene network analysis of differentially methylated genes was done using Ingenuity Pathway Analysis (Qiagen, Germantown, MD, USA). ToppGene Suite was used for functional gene ontology enrichment analysis(31). Molecular Function and Biological Process Gene Ontologies and KEGG Pathways were selected for enrichment. P-values were derived using a hypergeometric probability mass function and a Benjamini-Hochberg FDR-adjusted P-value cutoff of < 0.05 was used as a threshold of significance. Ontologies and pathways had to have a minimum membership of 3 genes and maximum of 2000 genes to be included.

Interferon-regulated genes were identified using the differentially methylated gene set as input for Interferome (v.2.01)(32) limiting results to genes with an expression fold change of 1.5 or greater between type I interferon-treated and untreated samples using datasets derived from peripheral whole blood.

### Statistical Analysis

We used probe-wise linear regressions to detect CpG sites in our cohort that show methylation difference between African-American and European-American patients using the *limma* (v.3.42.2) package(33). Patient age and the top four medication components were adjusted for in each regression and an empirical Bayes moderated t-statistic and P-value calculated for each probe. CpG sites were considered to be significantly differentially methylated if they had a Benjamini-Hochberg FDR-adjusted P-value < 0.05 and were differentially methylated by at least 10% between African-American and European-American patients.

Methylation M-values from the initial time point samples (n = 53), sample genotypes (n = 53), sample age, the top four medication components, and top ten genotype principal components were used to build a linear model for detecting meQTLs using *MatrixEQTL* (v.2.3) in R(34). *Cis*-meQTLs were defined as CpG sites with methylation values associated with a SNP within a conservative 1000bp of the CpG dinucleotide. We used a Benjamini-Hochberg FDR-adjusted P-value cutoff of < 0.05 for significant associations.

Analysis of the association with SLEDAI score and DNA methylation in our longitudinal cohort (n = 93 African-American & 136 European-American patient samples) was performed by fitting a linear mixed model using the *lmerTest* (v.3.1-2)(35) and *MuMIn* (v.1.43.17) packages in R(36). A regression model was fit in a probe-wise manner for all samples in each ancestry group to allow detection of ancestry-specific associations.

Regression models were adjusted for age at sample collection as a fixed effect and SLEDAI score as the variable of interest. Repeated samples were grouped by patient which was accounted for as a random effect in the model. CpG methylation and SLEDAI score had a statistically significant association if they had a Benjamini-Hochberg FDR-adjusted P-value < 0.05 and a suggestive association with a Benjamini-Hochberg FDR-adjusted P-value < 0.10. The impact of adjusting for medication components was determined by comparing the fit of the previously specified mixed effect regression model with an extended model that includes additional fixed effects for the top four medication components of each lupus patient’s timepoint. A chi-square difference test for nested models was applied using the *anova* function in R to determine if model fit was improved. A P-value < 0.05 was considered statistically significant and indicates that the larger model has improved data fit.

Longitudinal analysis of nephritis in our cohort was performed by fitting a linear mixed model as above to each probe using methylation profiles for n = 11 lupus patients (n = 7 African-American and n = 4 European-American) at a timepoint with active nephritis (as defined by SLEDAI) and the nearest preceding or receding timepoint without nephritis after adjusting for the top four medication components, age, and ancestry as fixed effects. Sample pairs were included as a random effect. A Benjamini-Hochberg FDR-adjusted P-value threshold of < 0.05 was used to identify statistically significant associations.

Two-group testing of mean ages between ancestry groups was done using a t-test. SLEDAI criteria and medication differences were compared using Fisher’s exact test. Comparing allelic proportions between ancestry groups was done using a two-proportion z-test. All P-values were two-tailed and a significance threshold of P < 0.05 was used.

## Results

### Longitudinal analysis of DNA methylation over time and across disease activity levels in lupus patients

The Infinium MethylationEPIC array measures the methylation status of 866,836 methylation sites across the genome including 863,904 CpG and 2932 CNG sites (C, cytosine; N, any nucleotide; G, guanine)(37). After QC and technical probe masking, a total of 745,477 (86.0%) sites were retained for analysis. Heterogeneity in disease manifestations, patient genetic background, and the environment are all factors that complicate the understanding of lupus pathogenesis. Using repeated sampling of lupus patients followed longitudinally, we can account for these factors and detect novel changes in DNA methylation that are associated with disease activity over time. We followed a total of 54 lupus patients for up to 43 months and assessed genome-wide DNA methylation levels in neutrophils in a total of 229 patient samples. Our cohort included 22 African-American and 32 European-American lupus patients followed across 93 and 136 timepoints, respectively. We assessed correlation between DNA methylation changes in individual methylation sites across the genome with disease activity as measured by SLEDAI scores in each ancestry group. After removing CpG-SNP probes with a minor allele frequency > 1% to avoid a bias due to intra-ancestral allele frequency differences, we analyzed a total of 733,192 (84.6%) methylation sites.

In the African-American cohort we identified a total of eight CpG sites that met our suggestive FDR-adjusted P-value < 0.1 (**Figure 1a, Table 1**). Two sites cg26104306 (*SNX18*; FDR-adjusted P-value = 3.38E-02) and cg06708913 (FDR-adjusted P-value = 3.43E-02) were significantly associated with changing disease activity in our cohort (**Figure 1b and 1c**). Cg26104306 shows stark demethylation with increasing disease activity compared to our European-American patients which showed very little methylation change across time and disease activity. Similarly, cg06708913 shows a much higher rate of increasing methylation with disease activity in African-American patients relative to European-American patients. The inclusion of the top four medication components as fixed effects did not improve the fit of our model for these two CpG sites (cg26104306 Chi-square P-value = 0.25 and cg06708913 Chi-square P-value = 0.83). Our European-American sample cohort analysis did not identify any CpG-SLEDAI score associations at either P-value threshold. Importantly, these data suggest that DNA methylation patterns defining lupus patients are largely stable over time and across disease activity.

**Figure 1:**
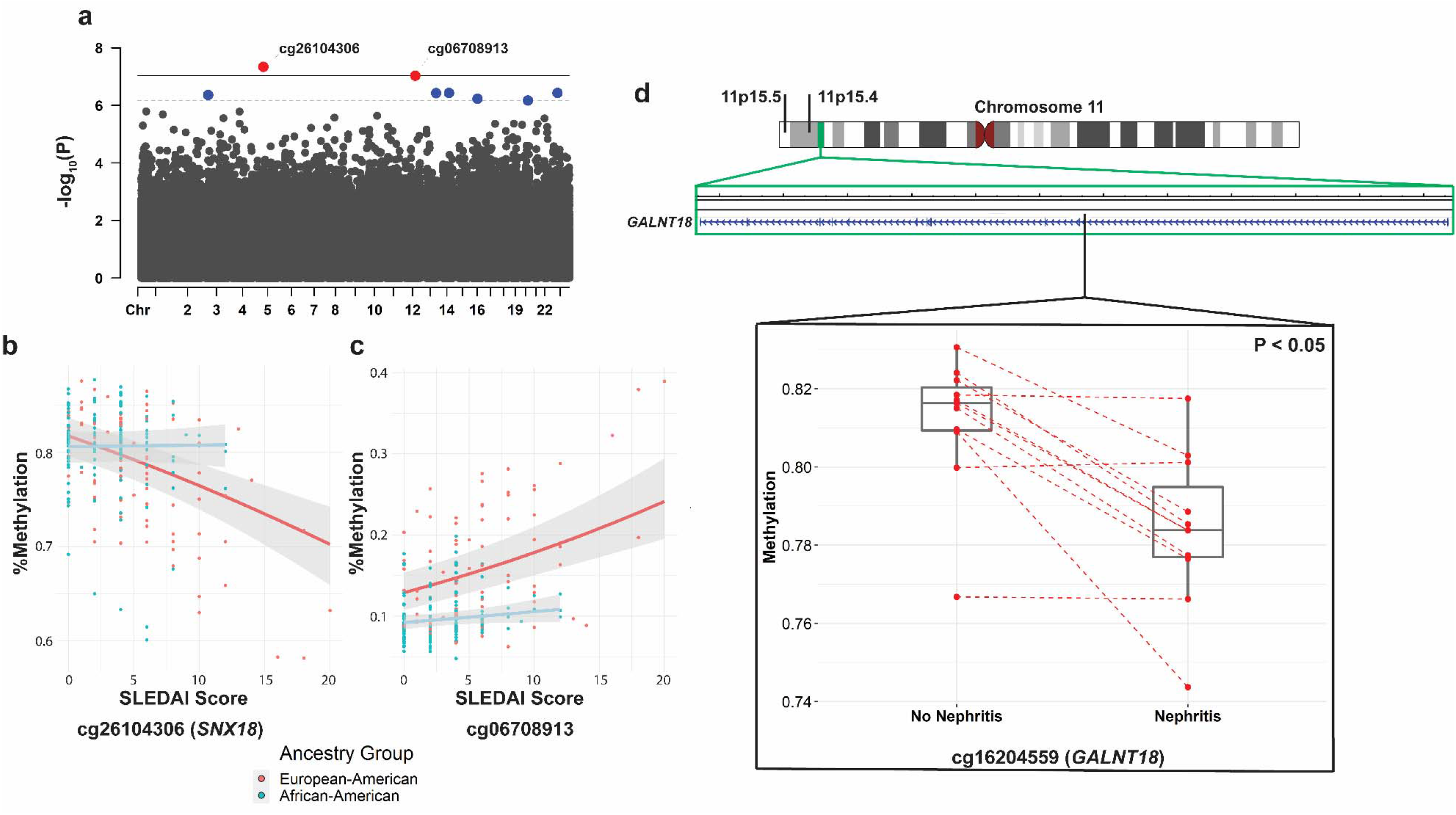
(a) A Manhattan plot depicting the significance of correlation between methylation levels of CpG sites and disease activity as measured using SLEDAI scores in African-American lupus patients. The red dots are CpG sites that meet the threshold for significance of FDR-adjusted P-value < 0.05 (bold line) and the blue dots are CpG sites that meet the suggestive threshold of FDR-adjusted P-value < 0.10 (dashed line). (b) Methylation status of cg26104306 (P = 4.61E-08; FDR-adjusted P = 3.38E-02) and cg08708913 (P = 9.35E-08; FDR-adjusted P = 3.43E-02) across SLEDAI scores for African-American (n = 93 samples; red dots/line) and European-American (n = 136 samples; blue dots/line) lupus patients. (d) Ideogram of chromosome 11 showing the location of 11p15.5 and 11p15.4 cytobands. Cg16204559 (black line) is within the body of *GALNT18* located in the 11p15.4 region (green box). Methylation profiles for n = 11 lupus patients (red dots (n = 7 African-American and n = 4 European-American) at a timepoint with active nephritis and the nearest preceding or receding timepoint without nephritis were compared after adjusting for medications, age, and ancestry group. Cg16204559 (*GALNT18*) was significantly demethylated (Mean β Nephritis: 78.4% and Mean β Non-nephritis: 81.2%; FDR-adjusted P = 0.048) with the occurrence of nephritis.

We next performed an analysis in a subset of patients who developed active lupus nephritis at any time point during our study and in whom a sample from at least one time point without evidence of lupus nephritis is available. After adjusting for medications, age, and ancestry we identified a single CpG site with a statistically significant relationship between DNA methylation levels and active nephritis in n = 11 lupus patients. DNA methylation levels in cg16204559, which is located within the gene *GALNT18*, are significantly reduced during active nephritis in lupus patients (**Supplementary Table 2, Figure 1d**).

### DNA methylation differences in neutrophils of African-American and European-American lupus patients

We then performed a differential DNA methylation analysis comparing lupus patients with African-American (n = 22) and European-American (n = 32) ancestry after adjusting for medication use and age. A multidimensional plot of the 5000 most variable CpG sites in these patients showed that methylation patterns tended to cluster by patient ancestry group (**Figure 2a**). African-American lupus patients in our cohort had more active disease compared to European-American lupus patients (SLEDAI 5.2±4.5 vs 2.9±3.2, respectively; P-value (t-test) = 0.03). Medication use at the initial timepoint were not significantly different between the ancestry groups (**Supplementary Table 1**). We identified 907 differentially methylated CpG sites using an FDR-adjusted P-value threshold of < 0.05 and a differential methylation between ancestry groups of at least 10% (**Figure 2b, Supplementary Table 3**). 487 (53.7%) of these sites were hypomethylated in African-American compared to European-American lupus patients and 420 (46.3%) were hypermethylated (**Figure 2c**). DNA methylation levels differed by 16.5% on average (SD:8.2%; range:10.0% - 57.9%) between ancestry groups. The hypomethylated and hypermethylated sites were associated with 391 and 316 genes, respectively. Hypomethylated genes showed enrichment for gene ontologies for granulocyte differentiation (GO:0030852; FDR-adjusted P-value = 2.23E-02 & GO:0030853; FDR-adjusted P-value = 3.20E-02), cell adhesion (GO:0007155; FDR-adjusted P-value = 1.26E-02 & GO:1903037; FDR-adjusted P-value = 3.41E-02), and toll-like receptor (TLR) signaling pathways (GO:0002224; FDR-adjusted P-value = 3.41E-02 & GO:0034121; FDR-adjusted P-value = 4.46E-02) (**Supplementary Table 4**). Hypermethylated genes were enriched for fewer ontologies representing primarily Rho guanine nucleotide exchange factor (GEF) protein activity (GO:0005089; FDR-adjusted P-value = 7.76E-03) and ATP binding (GO:0005524; FDR-adjusted P-value = 2.80E-02) (**Supplementary Table 5**).

**Figure 2:**
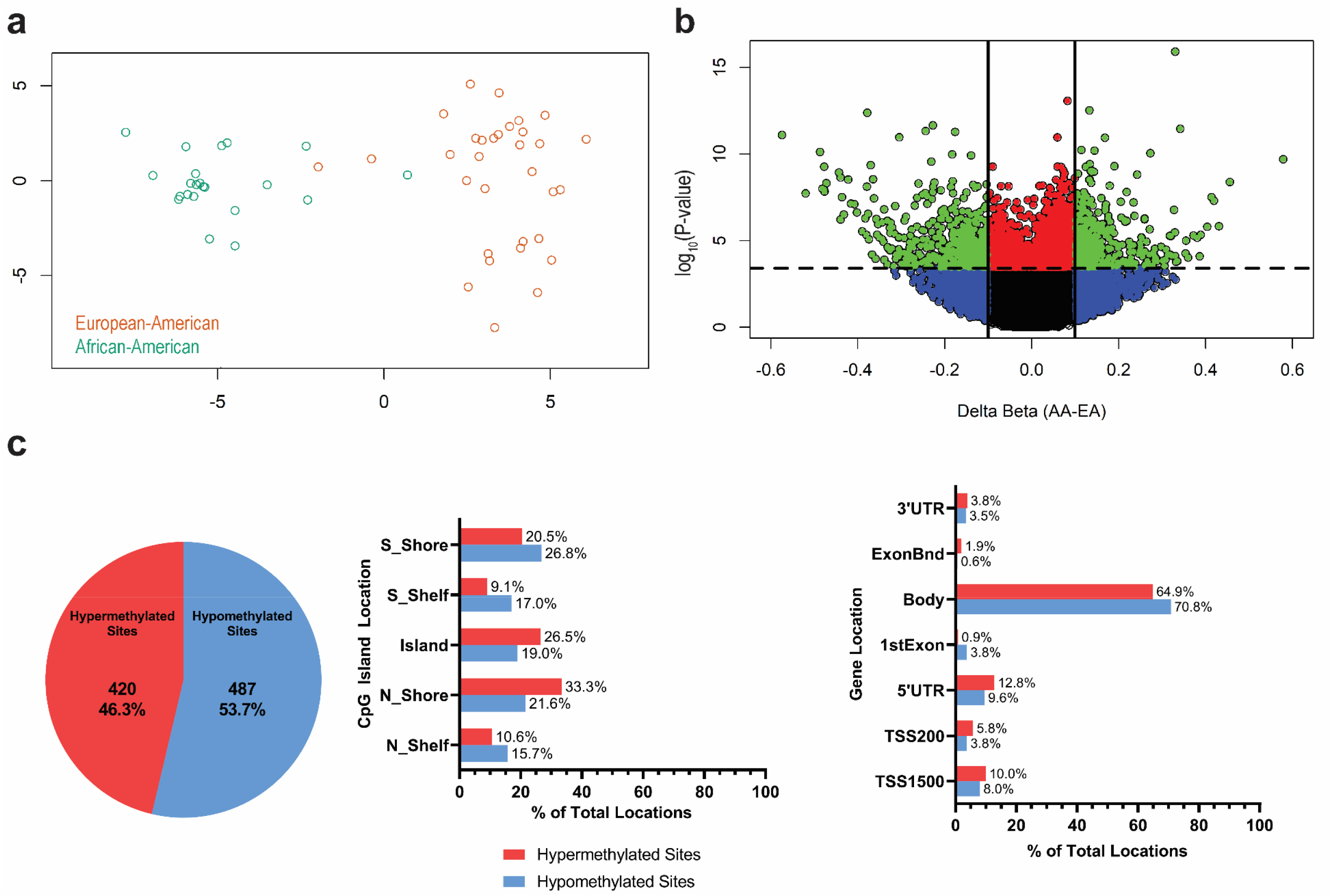
(a) Multidimensional scaling plot of top 5000 most variable CpG sites in African-American (n = 22; green circles) and European-American (n = 32; orange circles) lupus patients at initial sample collection. (b) Volcano plot of differentially methylated CpG sites between African-American (n = 22) and European-American (n = 32) lupus patients at initial sample collection. Each dot represents a CpG site (n = 745,477). Significantly differentially methylated sites (green) are differentially methylated by at least 10% between ancestry groups and with an FDR-adjusted P-value < 0.05 (n = 907). (c) Pie chart (left) showing the percent of sites hypermethylated (n = 420 (46.3%)) and hypomethylated (n = 487 (53.7%)) in African-American lupus patients compared to European-American controls. Barcharts showing the distribution of hypermethylated (red) and hypomethylated (blue) sites annotated to locations with CpG islands and genes (middle and left, respectively). S_Shore: South Shore; S_Shelf: South Shelf; N_Shore: North Shore; N_Shelf: North Shelf. 3’UTR: 3’ Untranslated Region; ExonBnd: Exon Boundary; 5’UTR: 5’ Untranslated Region; TSS200: 200bp upstream of Transcription Start Site; TSS1500: 1500bp upstream of Transcription Start Site.

### Methylation quantitative trait loci analysis

We next identified associations between DNA methylation and genotype in our cross-sectional cohort of lupus patients (n = 53) after controlling for age, medications, and genetic background. *Cis*-meQTLs in our cohort (**Figure 3a, Supplementary Table 6**) were defined using a conservative range of 1000bp to focus on localized effects. We identified a total of 8855 pairs of CpG sites and SNPs with an FDR-adjusted P-value <0.05. These meQTL pairs represented 7614 (86.0%) unique methylation sites and 7094 (80.1%) unique polymorphisms. 7269 (82.1%) of meQTLs did not contain CpG-SNPs. Gene set enrichment analysis of the 3871 unique genes associated with the CpG sites revealed numerous ontologies and pathways. The most significantly enriched included ontologies and pathways for cell and biological adhesion (GO:0007155; FDR-adjusted P-value = 6.43E-20, GO:0022610; FDR-adjusted P-value = 6.43E-20, & KEGG:83069; FDR-adjusted P-value = 5.29E-04) and calcium ion binding and signaling pathways (GO:0005509; FDR-adjusted P-value = 1.46E-07 & KEGG:83050; FDR-adjusted P-value = 9.58E-04) (**Figure 3b, Supplementary Table 7**). The meQTLs revealed in our study are, at least in part, responsible for the observed DNA methylation differences between African-American and European-American patients. Indeed, of the 907 differentially methylated CpG sites in our cohort, 142 (15.7%) were also meQTLs (**Figure 3a, Supplementary Table 8**). These included sites associated with *IL16* (cg02810829; Δβ = −0.23) and an meQTL associated with the triggering receptor expressed on myeloid cells (TREM)-like 4 gene *TREML4* (cg25555787; Δβ = −0.20). Cg25555787 had one of the strongest meQTL associations in this study (rs9369265 meQTL R^2^ = 0.91) (**Figure 3c**). We identified 1586 (17.9%) meQTLs that were tagged as including CpG-SNPs and comprised many of the strongest methylation-genotype associations (**Supplementary Table 6**).

**Figure 3.**
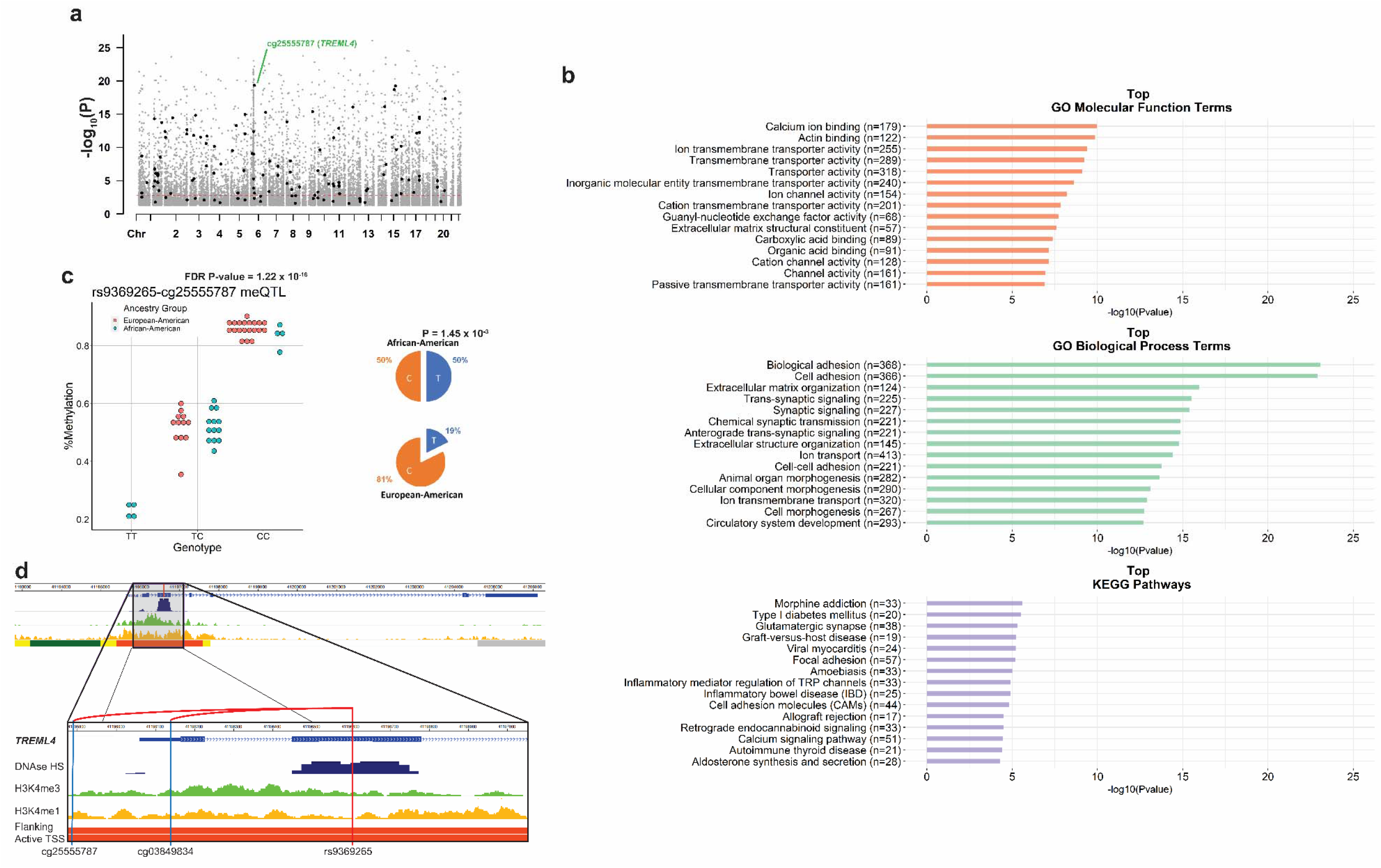
(a) A Manhattan plot of CpG site locations (black and grey dots) in *cis*-meQTL pairs with a SNP in our lupus cohort. Black dots represent CpG sites in non-CpG-SNP *cis*-meQTL pairs that had a significantly different average methylation between African-American and European-American patients (FDR-adjusted P-value < 0.05). The red dashed line represents an approximate FDR-adjusted P-value threshold of 0.05 for all *cis*-meQTLs across the entire genome. (b) Enrichment of gene ontologies and pathways of all *cis*-meQTLs across the genome (FDR-adjusted P-value < 0.05). Barcharts show the most significant molecular function (orange) and biological process (green) gene ontology terms, and KEGG pathways (purple) by −log_10_(P-value). (c) Rs9369265 is significantly associated with the methylation status of cg25555787 (FDR-adjusted P-value = 1.22E-16). The allele frequency of rs9369265 significantly differed between European-American (n = 32) and African-American (n = 21) lupus patients by 31% with the T allele associated with lower DNA methylation (P = 1.45E-3). (d) Rs9369265 is an exonic SNP in *TREML4* and is significantly associated with the methylation status of two CpG sites upstream of the transcription start site of *TREML4* (cg25555787 & cg03849834) (hg19). This region has epigenetic marks including DNase hypersensitivity (DNase HS), histone 3 lysine 4 mono-(H3K4me1) and tri-methylation (H3K4me3) and is labelled as an enhancer region for *TREML4* (“Flanking Active TSS”; orange bar) in primary human neutrophils. Data for figure panel (d) were generated using the WashU Epigenome Browser using ENCODE and Epigenome Roadmap ChromHMM data tracks from peripheral primary human neutrophils (E030).

### Methylation quantitative trait loci involving susceptibility and type I interferon genes for lupus

Comparing methylation site-associated genes in meQTL pairs with previously identified lupus susceptibility loci from genome-wide association studies(38-42) we identified 79 meQTL pairs (8.7%) in 28 lupus susceptibility genes (**Supplementary Table 9**). These included interferon regulatory factors *IRF7* and *IRF8* and *STAT4* which are involved in the type I interferon response. To identify type I interferon regulated genes that are associated with meQTLs in our cohort, we compared our meQTL-associated genes with the genes included in the Interferome (v.2.01) database(32). 64 of the 3871 unique genes (1.7%) associated with methylation sites in meQTLs were identified as type I interferon regulated genes (**Supplementary Table 10**).

## Discussion

Neutrophils are the most numerous cells in circulating blood and are early responders to inflammatory events throughout the body. They play an important role in entering sites of infection to identify pathogens through a variety of receptors, destroying pathogens,and secreting inflammatory signals to mobilize the immune system in response(43). Their primary methods of destroying pathogens include phagocytosis, production of reactive oxygen species, release of granules containing antimicrobial enzymes, and the release of NETs which physically bind and expose pathogens to antimicrobial proteins(43). In lupus, neutrophils display several abnormal phenotypes including enhanced apoptosis, increased NETosis after type I interferon priming, and impaired phagocytosis(44). Our prior work has found that lupus neutrophils display a DNA methylation signature common to other immune cell types, primarily demethylation of type I interferon response genes(12).

We interrogated the DNA methylome of neutrophils in a cohort of lupus patients followed longitudinally for about four years across 229 timepoints to assess DNA methylation changes over time and across different levels of disease activity. We showed that the DNA methylome is largely stable over time and across disease activity in lupus patients. We identified two CpG sites (cg26104306 and cg06708913) with DNA methylation levels that significantly correlated with disease activity. These correlations were detected in African-American but not European-American lupus patients.

Cg26104306 lies 745bp upstream of the transcription start site of the gene *SNX18* which encodes the sorting nexin 18 protein SNX18 (**Figure 1b**). It is located on the 5’ north shore of a CpG island (chr5:54517549-54519476 (hg38)) that overlaps the *SNX18* promoter region. Methylated CpG islands are typically indications of silenced gene promoters in somatic cells and hypomethylation suggests disease-associated disruption in this silencing. SNX18 localizes to the plasma membrane of cells and plays a functional role in endocytosis and autophagosome formation in cells(45, 46).

Cg06708913 overlapped a long non-coding RNA AC009522.1 and is proximal to an enhancer-like region denoted by transcriptionally permissive DNase hypersensitivity and increased H3K27ac modifications(47) (**Figure 1c**). One CpG site that reached suggestive significance for correlation with disease activity in African-American lupus patients, cg24682077 (FDR-adjusted P-values = 5.35E-02), is associated with FYVE, RhoGEF And PH domain containing 1 gene *FGD1*. FGD1 interacts with Rho GTPase Cdc42 which regulates neutrophil motility in response to extracellular signals(48). Cg24682077 is located 39bp downstream of the transcription start site of *FGD1* and within a promoter-associated CpG island.

The small number of CpG sites that change methylation levels with disease activity in our longitudinal study suggests that DNA methylation levels are stable in lupus neutrophils over time and across different disease activity levels. An inception study of lupus patients across time is necessary to detect DNA methylation biomarkers that indicate the onset of disease. A larger cohort size may bring more of these sites beyond the significance threshold or reveal novel associations in other ancestry groups. These associations will require replication to be confirmed but serve as indicators that novel disease-associated loci can be detected in longitudinal data from lupus patients. They also demonstrate that accounting for genetic ancestry in lupus studies can reveal novel associations.

Lupus nephritis is one of the most severe manifestation of lupus that can lead to chronic kidney damage and renal failure. We compared two timepoints from lupus patients with samples collected with and without nephritis in the same patient and adjusted DNA methylation changes for medication use, age, and ethnicity. A single methylation site, cg16204559, passed our FDR significance threshold corrected for multiple testing. Cg16204559 (chr11:11451256-11451258 (hg38)) is in the 11p15.4 cytoband within an intron of the gene *GALNT18* which encodes the polypeptide N-acetylgalactosaminyltransferase 18 protein. 11p15.4 is adjacent to 11p15.5 which has previously been identified as the location of the Systemic Lupus Erythematosus Nephritis 3 (*SLEN3*) locus. This locus is near the p telomere of chromosome 11 and was identified as a susceptibility locus for lupus using genetic linkage in multiplexed pedigrees of African-American ancestry that included lupus patients with nephritis(49). Understanding the biological role of this demethylation in lupus nephritis will require further investigation. Our study reveals the value of using longitudinal epigenetic studies to identify novel DNA methylation changes that could provide insight for specific disease manifestations.

Ancestry-specific DNA methylation differences and meQTL analyses showed a significant enrichment in Rho GEF pathways. GEFs are proteins that catalyze the cycling of GDP/GTP binding in Rho GTPases which results in their activation(50, 51). Rho GTPase activity regulates neutrophil function by controlling cytoskeletal arrangements in response to activation of signaling pathways(52). They regulate reactive oxygen species production, endothelial adhesion and transmigration, and production of neutrophil extracellular traps (NETs)(52). We used network analysis to further characterize ancestry-specific differential DNA methylation in lupus.

**Supplementary Figure 3** shows a gene network that is centered around the transcription factor complex nuclear factor kappa B (NFκB). NFκB is activated through degradation of inhibitory proteins in response to inflammatory signaling, such as TLR engagement, where it then translocates to the nucleus (53). There it coordinates the expression of proinflammatory gene programs in neutrophils that delay apoptosis, promotes production of proinflammatory cytokines, increases cell adhesion, and NETosis when cells are sufficiently activated(54). Resting human neutrophils tightly regulate NFκB activation through high levels of nuclear IκBα that is rapidly degraded upon proinflammatory stimulation(55). The gene *IKBKB* (cg20242624; Δβ = −0.11) encodes the inhibitor of NFκB kinase subunit beta protein (IKKβ) which is part of the IκB kinase (IKK) complex which is required for activation and nuclear translocation of NFκB by phosphorylation of the NFκB inhibitory subunit IκBα(56). *BCL10* (cg17322118; Δβ = −0.18) encodes the B-cell lymphoma/leukemia 10 protein BCL10 which is also an activator of NFκB through ubiquitination of the IKK subunit protein IKKγ(57).

Hypomethylation of these genes in neutrophils may reflect an increased response to inflammatory stimuli that promotes tissue invasion and inflammatory damage. Differentially methylated genes involved in regulating the type I interferon response were also present (**Supplementary Figure 4**). In particular *IRF7* (cg08926253; Δβ = −0.14 & cg22016995; Δβ = 0.13) was significantly hypomethylated similar to what we previously observed in naïve CD4+ T cells of African-American lupus patients and the neutrophils of lupus patients compared to healthy controls(12, 58). *IFI44*, also a type I interferon response gene was also significantly hypomethylated in African-American patients (cg01079652; Δβ = −0.23). We compared differentially hypomethylated genes in African-American lupus patients to the Interferome (v.2.01) database(32) to identify other type I interferon-regulated genes (**Supplementary Table 10**). Of interest, the cytokine gene *IL16* was hypomethylated in African-American patients (cg02810829; Δβ= −0.23) and had a modest association in an meQTL pair (rs35130261 meQTL R^2^ = 0.68; ΔMAF = 0.33). IL-16 is a chemoattractant cytokine that induces infiltration of T cells, macrophages, and eosinophils into sites of inflammation, and promotes pro-inflammatory cytokine release by monocytes *in vitro*(59). It also promotes IL-2 receptor expression on the surface of CD4+ T cells, enhancing IL-2 activity(60) and the migration and expansion of regulatory T cells in sites of inflammation(61). A recent study observed that neutrophils produce and store inactive pro-IL-16 in the cytosol which is released and activated by caspase-3 upon secondary necrosis(62). Increased circulating IL-16 levels in lupus patients is associated with more severe disease(63, 64), and primary neutrophils of lupus patients more readily undergo apoptosis and increased secondary necrosis with reduced clearance of apoptotic material(65). This suggests that hypomethylation of *IL16* (in part related to meQTLs) could promote an exaggerated inflammatory response upon neutrophil secondary necrosis in lupus patients.

A demonstration of the mechanism underlying meQTL associations can be seen in two of the strongest meQTL pairs, cg25555787 (*TREML4*; rs9369265 meQTL R^2^ = 0.91; ΔMAF = 0.31) (**Figure 3c**) and cg03849834 (*TREML4*; rs9369265 meQTL R^2^ = 0.81;ΔMAF = 0.31). Functionally, TREML4 has previously been identified as playing an important role in modulating the response to TLR7 signaling when bound to single-stranded RNA and TLR9 binding to unmethylated CpG-DNA(66). Rs9369265 lies in the second exon of *TREML4* within an active region flanking the transcription starts site of *TREML4* in neutrophils (**Figure 3d**). Rs9369265 genotype is also significantly associated with the expression of *TREML4* in whole blood (Gene-Tissue Expression Portal P-value = 1.2E-163), with the C allele associated with reduced expression and increased DNA methylation in our data. The presence of H3K4me3 peaks in this region and DNase accessibility suggest this is an important regulatory region for controlling *TREML4* expression as a promoter. A reduction in DNA methylation corresponds with an increase in H3K4me3 and promoter activity(67). The ligand for *TREML4* is unknown, but it readily binds to dead and dying cells(68). Reduced clearance of necrotic material in lupus patients might provide more stimulation to TREML4 and TLRs, promoting the exaggerated type I interferon response seen in lupus patients and contributing to the development of renal disease in lupus(69). Indeed, it has been observed that lupus-prone MRL*/lpr* mice have higher survival, produce fewer dsDNA autoantibodies and develop less renal damage when *Treml4* is knocked out(66). Neutrophils from *Treml4*^*-/-*^ mice show reduced expression of *Cxcl2* which is a potent neutrophil chemoattractant, but unimpaired motility and phagocytosis(66). The higher frequency of the T allele in our African-American lupus patients that correlates with increased *TREML4* expression suggests potential for a more robust response to TLR stimulation. This is supported by the observation of increased expression of the proinflammatory cytokines interferon alpha and TNF alpha in the whole blood of female African-American lupus patients compared to female European-American patients (70). The mechanisms underlying the association between genotype and DNA methylation status will require further study to identify. Potential mechanisms could include an inherited haplotype tagged by rs9369265 that promotes or suppresses transcription factor accessibility and binding, which is reflected in a repressive epigenetic state represented by increased DNA methylation correlating with reduced gene expression. This effect could also extend to other myeloid cells that express *TREML4* including macrophages and dendritic cells which contribute to the proinflammatory response(66).

## Conclusion

For the first time, we have analyzed the association of DNA methylation with disease activity across time in the neutrophils of lupus patients. We identified two CpG sites unique to patients of African-American ancestry with methylated levels associated with disease activity in lupus. We also identified a single CpG site with an association among lupus patients with the development of active lupus nephritis. To further characterize the ancestry-related DNA methylation changes in our cohort we identified meQTLs that were differentially methylated between ancestry groups. Two genes, *TREML4* and *IL16* contained meQTLs and were also significantly hypomethylated in African-American lupus patients. Both genes play roles in promoting inflammatory response to TLR signaling and infiltration of peripheral immune cells into tissue. Our data suggest that genetic background explains in part ancestry-specific patterns of DNA methylation patterns which have a direct impact on disease severity in lupus patients.

## Data Availability

All data reported in this paper are provided in the form of supplementary tables included with this submission.

## Acknowledgements

This work was supported by the National Institute of Allergy and Infectious Diseases of the National Institutes of Health grants number R01AI097134 and U19AI110502, and the Lupus Research Alliance.

## Figure legends

**Supplementary Figure 1:**
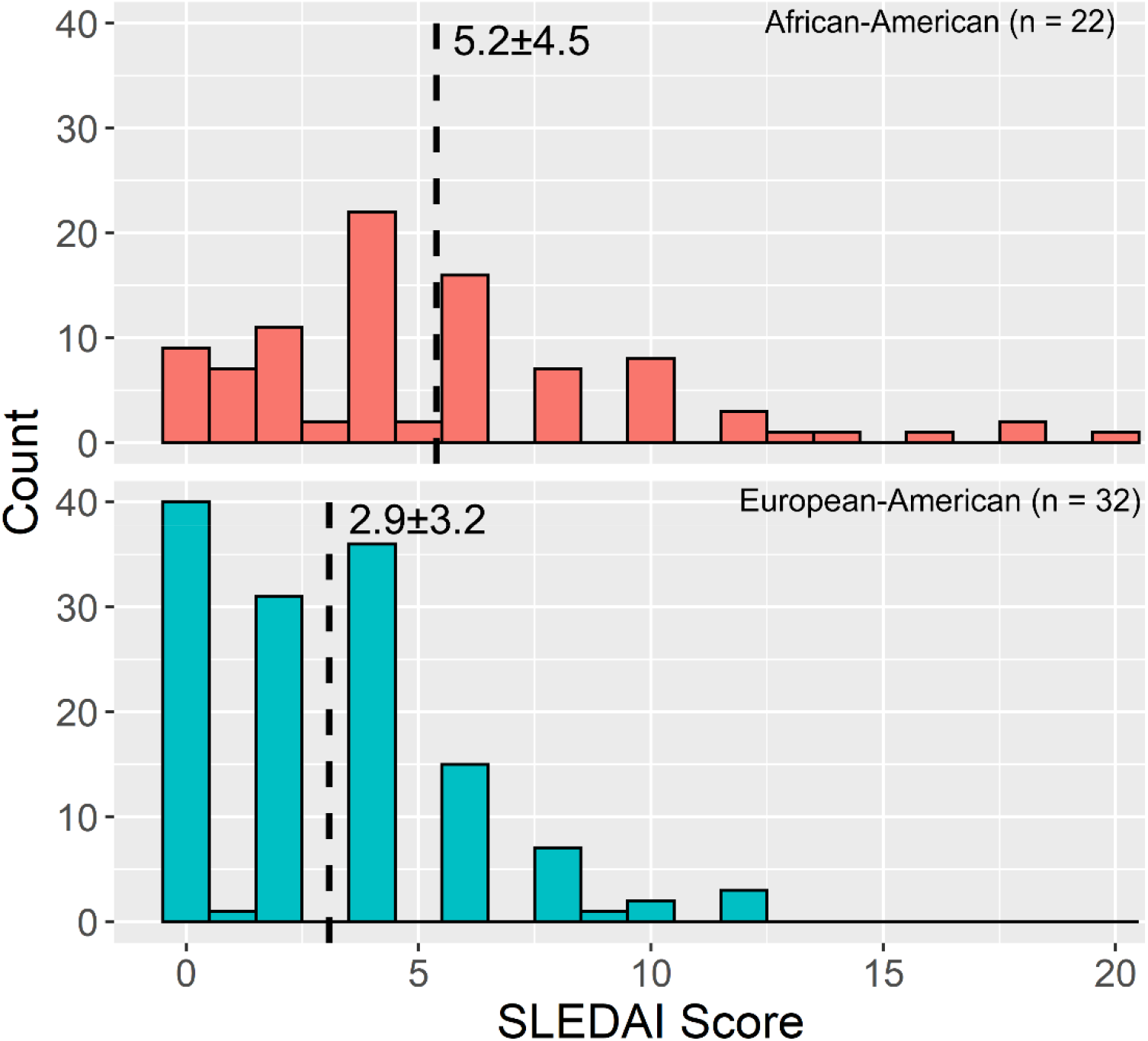
SLEDAI score distribution in African-American and European-American lupus patients at initial sample collection.

**Supplementary Figure 2:**
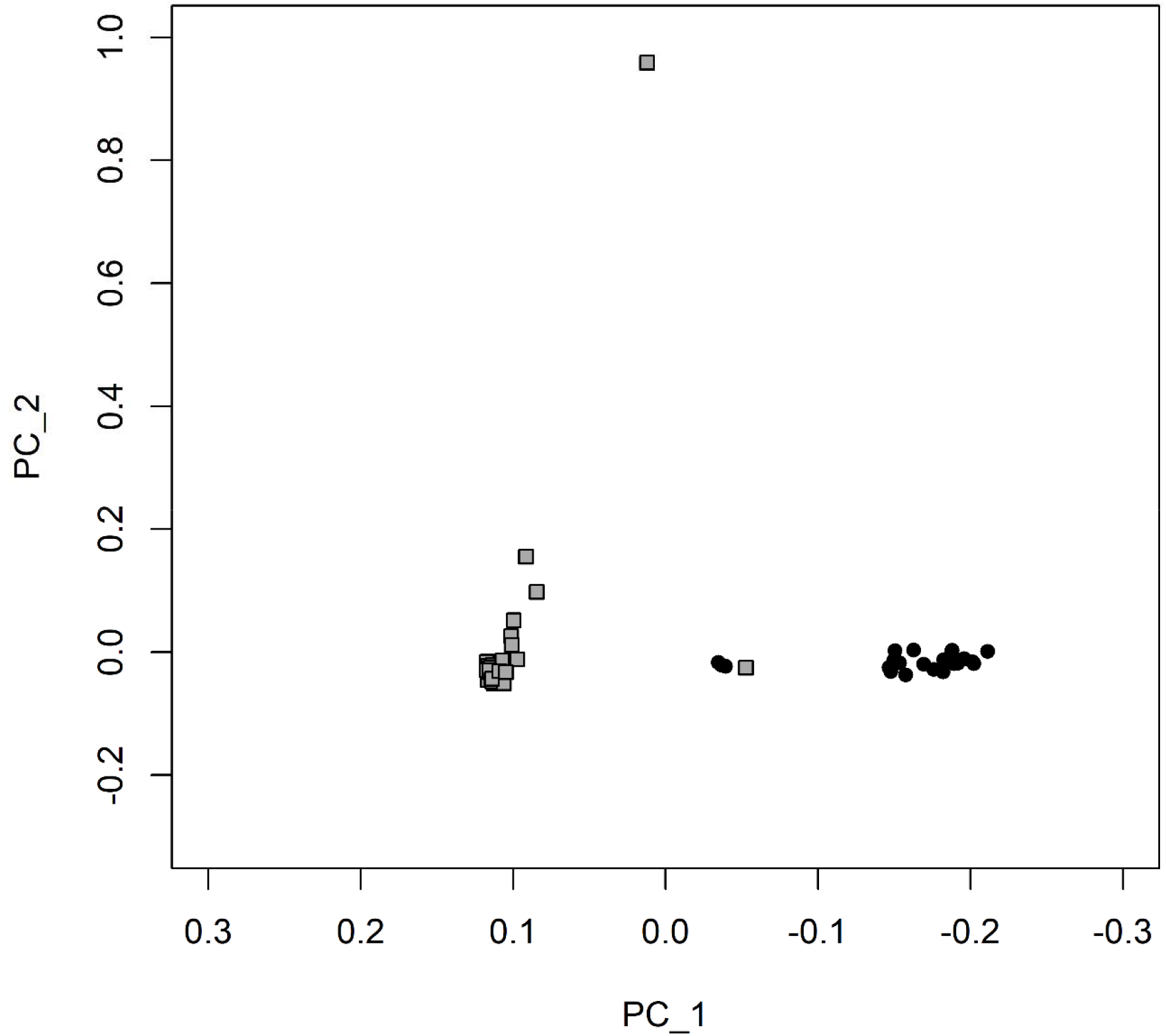
Dot plot of top two genotype principal components. Grey squares represent European-American lupus patients and black dots represent African-American lupus patients.

**Supplemental Figure 3.**
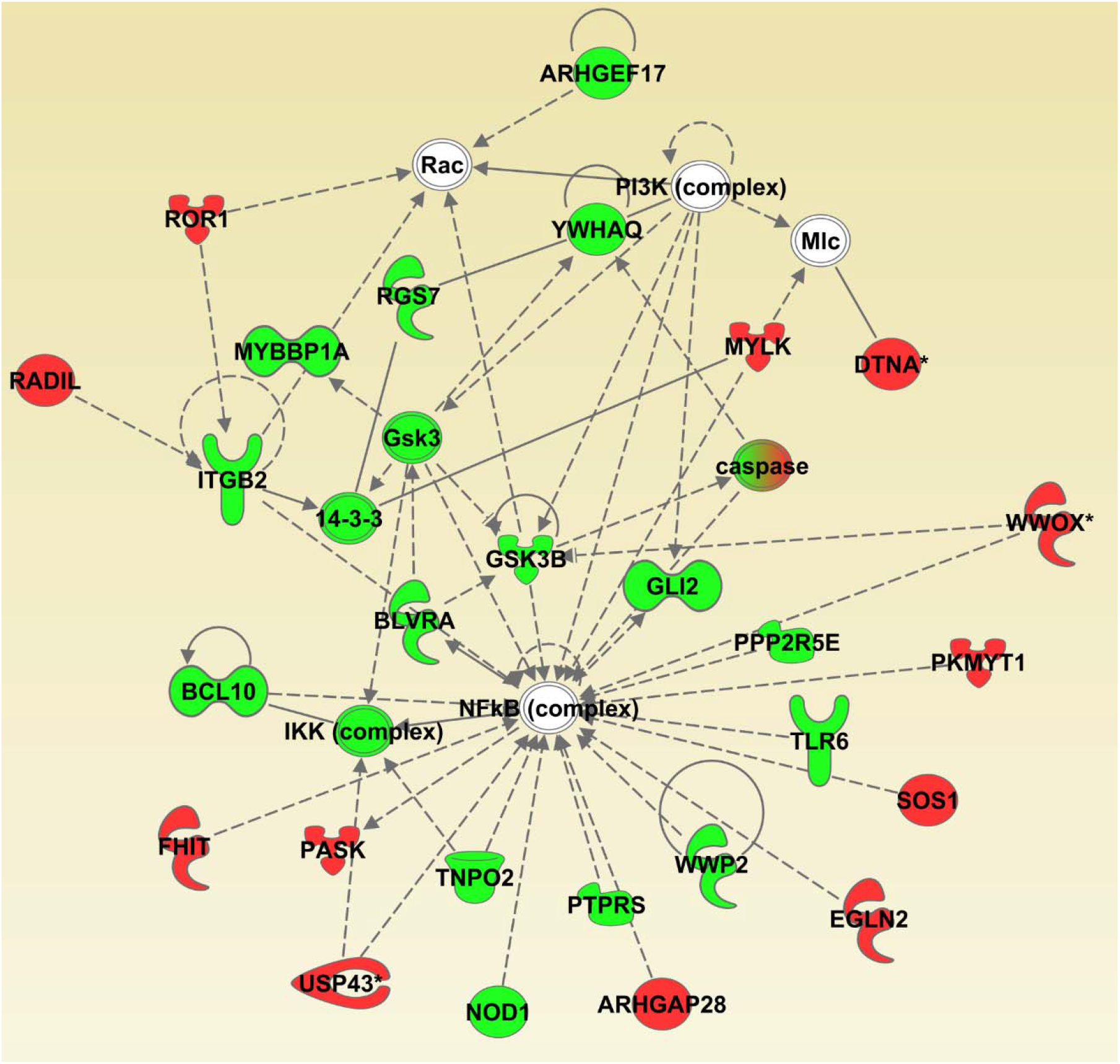
Networks of genes with hypomethylated (green shapes) and hypermethylated (red shapes) CpG sites in African-American compared to European-American lupus patients. White shapes represent genes included in the network by IPA through their relationship to the input genes. Green-red shapes included both hypo- and hypermethylated CpG sites. Dashed and solid lines represent indirect and direct interactions, respectively.

**Supplemental Figure 4:**
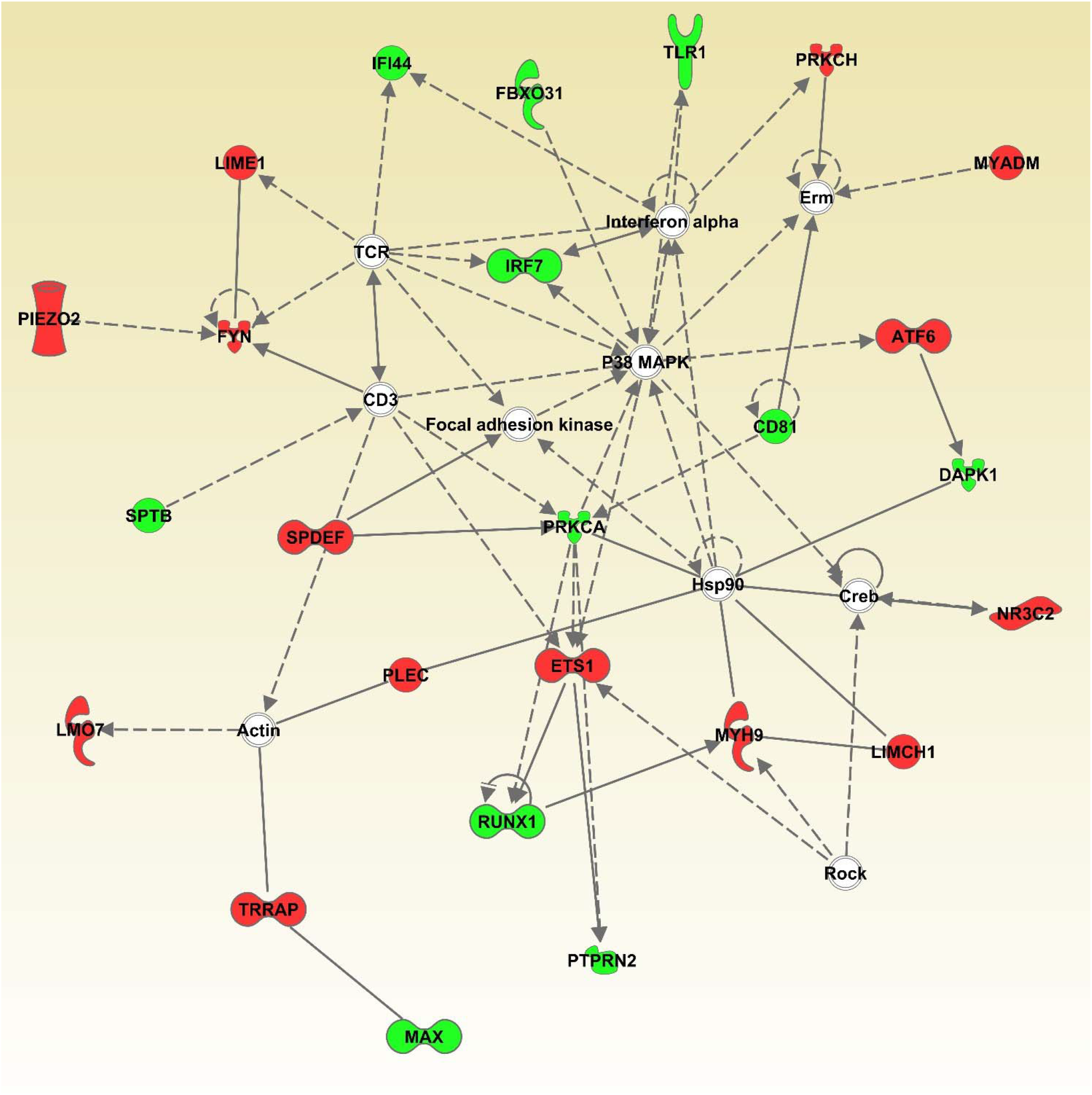
Networks of genes with hypomethylated (green shapes) and hypermethylated (red shapes) CpG sites in African-American compared to European-American lupus patients. White shapes represent genes included in the network by IPA through their relationship to the input genes. Dashed and solid lines represent indirect and direct interactions, respectively.

